# Menarche, pubertal timing and the brain: female-specific patterns of brain maturation beyond age-related development

**DOI:** 10.1101/2023.08.31.23294880

**Authors:** Nina Gottschewsky, Dominik Kraft, Tobias Kaufmann

## Abstract

**Background:** Puberty depicts a period of profound and multifactorial changes ranging from social to biological factors. While brain development in youths has been studied mostly from an age perspective, recent evidence suggests that pubertal measures may be more sensitive to study adolescent neurodevelopment, however, studies on pubertal timing in relation to brain development are still scarce.

**Methods:** We investigated if pre- vs. post-menarche status can be classified using machine learning on cortical and subcortical structural magnetic resonance imaging (MRI) data from strictly age-matched adolescent females from the Adolescent Brain Cognitive Development (ABCD) cohort. For comparison of the identified menarche-related patterns to age-related patterns of neurodevelopment, we trained a brain age prediction model on data from the Philadelphia Neurodevelopmental Cohort and applied it to the same ABCD data, yielding differences between predicted and chronological age referred to as brain age gaps. We tested the sensitivity of both these frameworks to measures of pubertal maturation, specifically age at menarche and puberty status.

**Results:** The machine learning model achieved moderate but statistically significant accuracy in the menarche classification task, yielding for each subject a class probability ranging from 0 (pre-) to 1 (post-menarche). Comparison to brain age predictions revealed shared and distinct patterns of neurodevelopment captured by both approaches. Continuous menarche class probabilities were positively associated with brain age gaps, but only the menarche class probabilities – not the brain age gaps – were associated with age at menarche.

**Conclusions:** This study demonstrates the use of a machine learning model to classify menarche status from structural MRI data while accounting for age-related neurodevelopment. Given its sensitivity towards measures of puberty timing, our work suggests that menarche class probabilities may be developed toward an objective brain-based marker of pubertal development.

**Highlights:**

✓ We classified pre- vs. post-menarche status in adolescent females from structural brain imaging data
✓ We compared class probabilities to brain-age predictions to disentangle puberty- vs. age-related patterns of brain development
✓ The derived continuous brain-based menarche class probabilities captured shared but also unique variations of adolescent neurodevelopment, and were associated with pubertal timing and status

**Plain English Summary:** Puberty is a period of substantial changes in the life of youths, and these include profound brain changes. Most studies have investigated age related changes in brain development, recent work however suggests that looking at brain development through the lens of pubertal development can provide additional insights beyond age effects. We here analyzed brain imaging data from a group of same-aged adolescent girls from the Adolescent Brain Cognitive Development study. Our goal was to investigate if we could determine from brain images whether a girl had started her menstrual period (menarche) or not, and we used machine learning to classify between them. This machine learning model does not just return a “yes/no” decision, but also returns a number between 0 and 1 indicating a probability to be pre- (0) or post- (1) menarche. To rule out, that our approach only maps age-related development, we selected a strictly age-matched sample of girls and compared our classification model to a brain age model trained on independent individuals. Our model classified between pre- and post-menarche with moderate accuracy. The obtained class probability was partly related to age-related brain development, but only the probability was significantly associated with pubertal timing (age at menarche). In summary, our study uses a machine learning model to estimate whether a girl has reached menarche based on her brain structure. This approach offers new insights into the connection between puberty and brain development and might serve as an objective way to assess pubertal timing from imaging data.

## Background

Adolescence is a time of profound changes to the body and the brain, with substantial impact on an individual’s behaviour, emotions, and self-perception, among other things [1]. This transition includes puberty, the time period during which an individual acquires the capability for sexual reproduction [2]. The latter is characterised by an interplay of gonadotropin-releasing hormone, gonadotropins such as follicle-stimulating hormone and luteinizing hormone, and sex hormones such as androgens, estrogens and progesterones. Together, they not only drive changes of the body, but also directly act on the brain [3]. Studies using magnetic resonance imaging (MRI) to investigate human brain anatomy have illustrated that the brain undergoes significant changes during adolescence described by a complex yet orchestrated interplay of progressive (e.g., myelination) and regressive (e.g., pruning) neuronal processes [4]. While brain development in youths has been commonly investigated through the lens of age-related brain maturation, there has been an increasing number of studies focusing on neurodevelopment mediated by pubertal processes in youth [5–7]. These studies suggest that puberty-related brain development cannot be simply explained by age trajectories but rather goes beyond the effects of growing older [7–9] and consequently that pubertal development may thus be a more sensitive measure to study neurodevelopment in youths as compared to age [6].

A recent systematic review on the relationship between pubertal and structural brain development in human adolescents describes brain wide reductions in cortical grey matter thickness and volume associated with progressed pubertal maturation from both cross-sectional and longitudinal studies [7]. Findings suggest that these effects are global across the brain with frontal regions showing the most pronounced effects [7,10]. Alongside cortical changes, advanced pubertal maturation is also associated with subcortical brain development, in particular the amygdala and hippocampus [11]. Across studies these effects are subject to sex differences, which not only manifest in varying effect sizes but also sometimes in opposing effect directions in males and females [5,12].

While methodological choices, such as accounting for age in statistical models, may factor into the diverging observations, these differences may also stem from variability that is inherent to pubertal maturation [13]. Although all individuals pass through the same pubertal stages, there is large variability regarding pubertal timing and the speed of progression [14,15]. Pubertal timing describes the time point at which an individual reaches certain pubertal milestones in comparison to their peers of the same age [16]. While pubertal timing appears to be highly heritable [17,18], recent evidence is also linking variation in pubertal timing to environmental factors, such as nutrition intake, socioeconomic status or obesity [14]. This malleability may consequently lead to pubertal onsets that deviate in their timing and individuals thus experiencing early or late pubertal onsets [5]. Interestingly these deviations in pubertal timing appear to be associated with physical and psychiatric health issues [5,19].

Many studies over the years have shown an association between pubertal timing and psychopathology [20–22]. In boys, evidence concerning the effect of pubertal timing on health risks is inconsistent and could be best described by the ‘off-time hypothesis’, that is either very early or very late onset [23]. In contrast, evidence for the association between health risk and puberty timing in girls has been well-replicated, converging on the so-called ‘early timing hypothesis’, which posits that early maturing girls (most often assessed using age at menarche as a proxy measure, i.e. age at which individuals experience their first menstruation) are more likely to experience adverse mental health outcomes than their on time and late maturing peers [24,25]. Therefore, pubertal timing and its malleability depict a critical tipping point which may set the course for later vulnerability and worse (mental) health outcome.

While most puberty-related imaging studies to date have focused on investigating the association between the brain and puberty status (i.e., the quantification of pubertal characteristics indicating a more or less advanced maturation akin to the transition through pubertal stages), imaging studies on pubertal timing – despite its importance for emerging (mental) health risks – are to the best of our knowledge scarce. The current study investigated the impact of pubertal timing on brain maturation, deploying age-matching to control for age-related neurodevelopment. Using structural imaging data from the Adolescent Brain Cognitive Development cohort (ABCD; [26]) we aimed at classifying pre- and post-menarcheal females using a machine learning model. To validate the sensitivity of our approach and to test the biological validity of the obtained class probabilities, we drew comparison to a brain age prediction framework, investigating to what extent both approaches capture the same or distinct neurodevelopmental variance in the female adolescent brain.

## Methods

### Sample Descriptions

ABCD: For the menarche classification and as the test sample for the age prediction model, we included data of N = 3248 female (henceforth referring to individuals assigned female at birth; mean age = 11.91 years, *SD* = 0.65) participants of the Adolescent Brain Cognitive Development study 2-year follow up data [26]. Study protocols have been approved by either local institutional review boards (IRB) or by reliance agreements with the central IRB at University of California. For each study participant, structural brain imaging features were obtained from the tabulated imaging data provided by the ABCD release 4.0 [27]. The 2-year follow up data was chosen because it offers the most balanced distribution of pre- and post-menarcheal individuals. Subjects with missing MRI or missing relevant demographic data were excluded. Furthermore, those who did not answer either ‘yes’ or ‘no’ to the question ‘Have you begun to menstruate (started to have your period)?’ from the ABCD Youth Pubertal Development Scale and Menstrual Cycle Survey History (PDMS) [28], or whose imaging data quality was deemed too low for inclusion by two ABCD researchers, were excluded. From the PDMS data we determined pubertal status ranging from prepubertal to postpubertal. In brief, we summed pubic hair growth and breast development scores and incorporated information about menarche and converted resulting score to a pubertal status category according to a scheme provided by the ABCD study (variable: pds_p_ss_female_category). Pubertal status was calculated from youth-reported as well as caregiver-reported data to account for differences in the perception of pubertal maturation.

PNC: We used data from N= 786 female participants (mean age = 15.25 years, SD = 3.65) of the Philadelphia Neurodevelopmental Cohort (PNC; [29]) as an independent training sample to derive an age prediction model. In PNC, all study procedures were approved by the respective institutional review boards. We processed the T1 MRI images using FreeSurfer (version 7.1.1) [30] and derived the same cortical and subcortical features as used for the ABCD cohort. Euler numbers were used as a proxy of image quality for quality control [31]. Subjects with missing MRI, missing demographic data, a Euler number more than three standard deviations below the mean, or those with a medical rating of 3 or higher (severe medical condition) were excluded.

### MRI data description

For each subject in both data sets we included a total number of 234 anatomical MRI features. Specifically, we used 30 subcortical features as well as, for each hemisphere, 34 volume, 34 thickness, and 34 area cortical features matching the Desikan-Killiany atlas [32] (see Supplementary Table S1). Of note, since ABCD data was acquired across 21 study sites, we performed batch harmonization with *neuroCombat* (v.0.2.12) [33] for each individual modality and training and testing sets independently.

### Statistical Analyses

All statistical analyses were performed in python (v.3.10.5) [34]. Basic data handling was performed with numpy (v.1.23.2) [35] and pandas (v.1.4.3) [36,37].

Menarche Classification: For the classification of pre- and post-menarche individuals in the ABCD sample, a linear discriminant analysis (LDA) classification model was trained using scikit-learn (version 1.1.1; [38]. For classification we split the full ABCD sample (N=3248 females) into a training and an independent test set by randomly sampling 20% of the data into the test set. The test sample consisted of N = 650 participants (pre-menarche: N = 419, mean age = 11.68 years, SD = 0.56; post-menarche: N = 231, mean age = 12.32 years, SD = 0.59). Furthermore, to avoid bias in the training process, propensity score matching [39] was performed in the training data to achieve equal distributions of age and MRI scanner in the pre- and post-menarche groups, as well as equal group sizes. After age-matching, there was no statistical age difference (two-sided independent samples t-test, p = 0.968) between the pre- and post-menarche groups in the training dataset (pre-menarche: N = 775, mean age = 12.09 years, SD = 0.58; post-menarche: N = 775, mean age = 12.09 years, SD = 0.58). Participants’ responses to the question ‘Have you begun to menstruate (started to have your period)?’ from the ABCD Youth Pubertal Development Scale and Menstrual Cycle Survey History (PDMS) were used as target labels for the classification algorithm. Responses were encoded numerically in the original survey as (4: Yes; 1: No) and relabelled to 0 and 1.

Train- and test set features were independently transformed into z-scores. Model tuning was performed via scikit-learn’s GridSearchCV. The hyperparameters explored in the grid search included the ’solver’ parameter with options [’svd’, ’lsqr’, ’eigen’] and the ’shrinkage’ parameter with values [None, ’auto’, 0, 0.1, 0.2, 0.3, 0.4, 0.5, 0.6, 0.7, 0.8, 0.9, 1]. The performance metric used for evaluation was accuracy and a 10-fold cross-validation approach was employed to assess the model’s performance during hyperparameter tuning. For the final model, an LDA model was fitted to the entire training dataset using the selected hyperparameters (solver: least squares solution, shrinkage: 0.8). A combined approach of 5-fold cross validation and a permutation test with 1000 permutations was employed to assess classifier performance. To assess the model’s performance on unseen data, the menarche status of participants from a held-out test sample of ABCD subjects was classified. An accuracy score and a confusion matrix were calculated. Furthermore, a permutation test with 1000 permutations was conducted to confirm that the accuracy of the classifier was significantly higher than chance. The estimated class probabilities of the withheld test sample were extracted from the LDA model to further assess the biological validity of the classification.

Brain age prediction: The python package of the XGBoost (v1.6.1) library was used [40] to predict chronological age in months from the same 235 sMRI features as those used in the menarche classification. Model tuning was again performed via scikit-learn’s GridSearchCV. The hyperparameters explored in the grid search were ’max_depth’: [3,6,9], ’max_leaves’: [0,2,5,10], ’learning_rate’: [0.001,0.01,0.1,0.5,1,3], ’min_child_weight’: [1,10,100] and ’n_estimators’: [100, 500, 1000]. The performance metric used for evaluation was mean squared error (mse) and a 5-fold cross-validation approach was employed to assess the model’s performance during hyperparameter tuning. The final model was fitted to the entire training dataset using the determined hyperparameters (’learning_rate’: 0.01, ’max_depth’: 6, ’max_leaves’: 0, ’min_child_weight’: 10, ‘subsample’: 0.5, ‘num_rounds’: 1000). Again, a combined approach of 5-fold cross validation and a permutation test with 1000 permutations was employed to assess classifier performance via root-MSE (RMSE) and mean absolute error (MAE). The model’s performance on unseen data was tested by applying it to the ABCD withheld test sample described above. The rmse and MAE were calculated and the brain age gap (difference between predicted brain age and chronological age; BAG) was calculated for further analysis.

### Association Analyses

Association analyses were performed with the ordinary least squares (OLS) regression function of the Python module *statsmodels* (v0.13.2) [41]. In line with previous studies [42,43], a residualised BAG was produced by regressing age and scanning site on BAG. Menarche class probabilities were residualised in the same way to account for age and scanning site. OLS regression was performed to test the association of residualised BAG and residualised menarche class probabilities. Finally, we tested for associations between age at menarche and residualised menarche class probabilities, as well as age at menarche and residualised BAG using OLS. Likewise, we tested for association between pubertal status and menarche class probability, as well as BAG, respectively. All association analyses were repeated accounting for potential effects of sociodemographic status (SES), body mass index (BMI) and race / ethnicity. In brief, BMI was calculated by averaging two height and weight measurements respectively and using the formula ‘height (lb) / height (in) x 703’. Ethnicity was encoded in 5 levels: White, Black, Hispanic, Asian, other (multiracial or ethnicity with too few members in the sample). Ordinal SES variables were transformed through rank-based inverse normal transformation and averaged, producing a single SES variable. Further details on the covariates and its calculations can be found in Kraft et al. [13].

## Results

We first tested if it was possible to classify from anatomical MRI between same-aged pre- and post-menarcheal girls. Our classifier trained in a sample of age-matched groups of females pre- and post-menarche performed with an accuracy of 60.00% in 5-fold cross validation (p < 0.001 across 1000 permutations; Fig. 1a). Applied to a held-out test set of 419 pre- and 231 post-menarcheal girls, the classifier performed equally well (61.23% accuracy, Fig. 1b, c).

**Figure 1:**
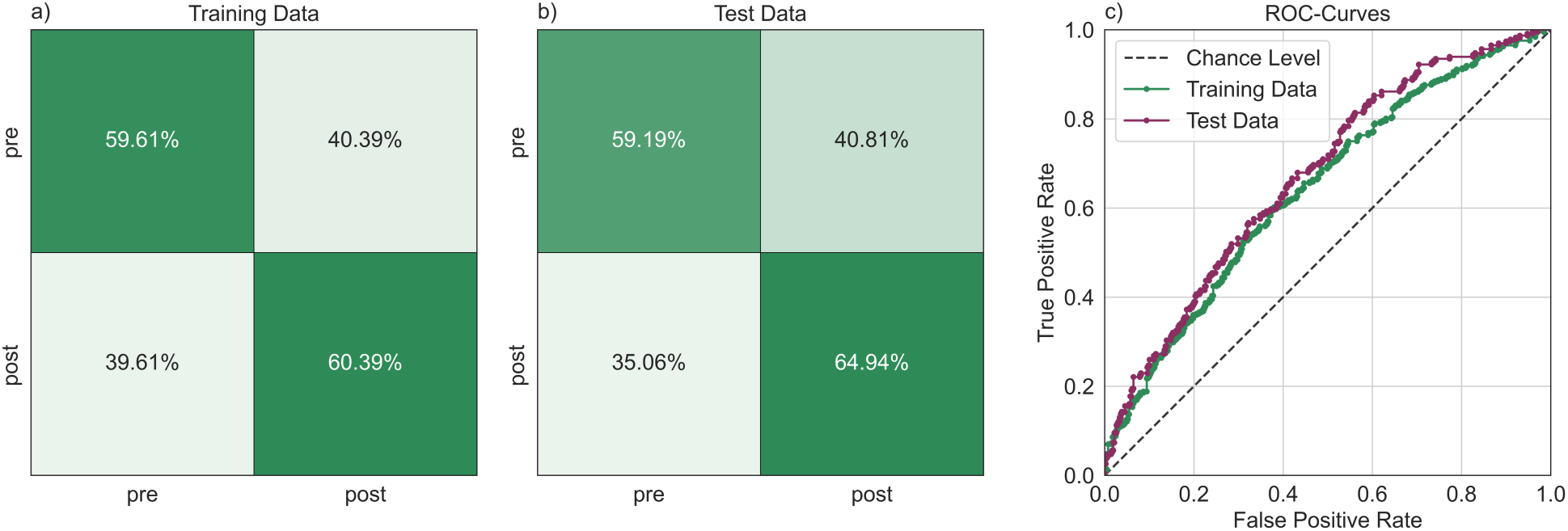
Menarche can be classified from brain imaging data. a) Confusion matrix of 5-fold cross validation in training data; b) Confusion matrix of classification model applied to hold-out test data; c) ROC curves of 5-fold cross validation and hold-out test data classification.

Figure 2a depicts the class probability obtained from the pre-/post-menarche classifier for each individual in the independent ABCD test sample. In an association analysis in the post-menarche group, we found an association of derived class probabilities and age at menarche. Specifically, individuals with an earlier menarche tend to be classified as post-menarche with a higher confidence (adj. *R*2 = 0.1.121, F(26, 188) = 2.129, t = -2.714, p = 0.007), supporting biological sensitivity of the class probabilities beyond the binary pre-/post distinction (Figure 2b). We furthermore found a positive association between menarche class probability and pubertal status (youth-reported PDMS: adj. *R*2 = 0.020, F(1, 629) = 13.94, t = 3.734, p < 0.001, caregiver-reported PDMS: : adj. *R*2 = 0.022, F(1, 616) = 14.76, t = 3.842, p < 0.001; Figure 3c), further corroborating biological sensitivity. Of note, both associations (age at menarche, puberty status) were diminished when incorporating BMI, SES and race/ethnicity as confound factors (Supplementary Table S2), highlighting the complex relationship between these factors, puberty and the brain.

**Figure 2:**
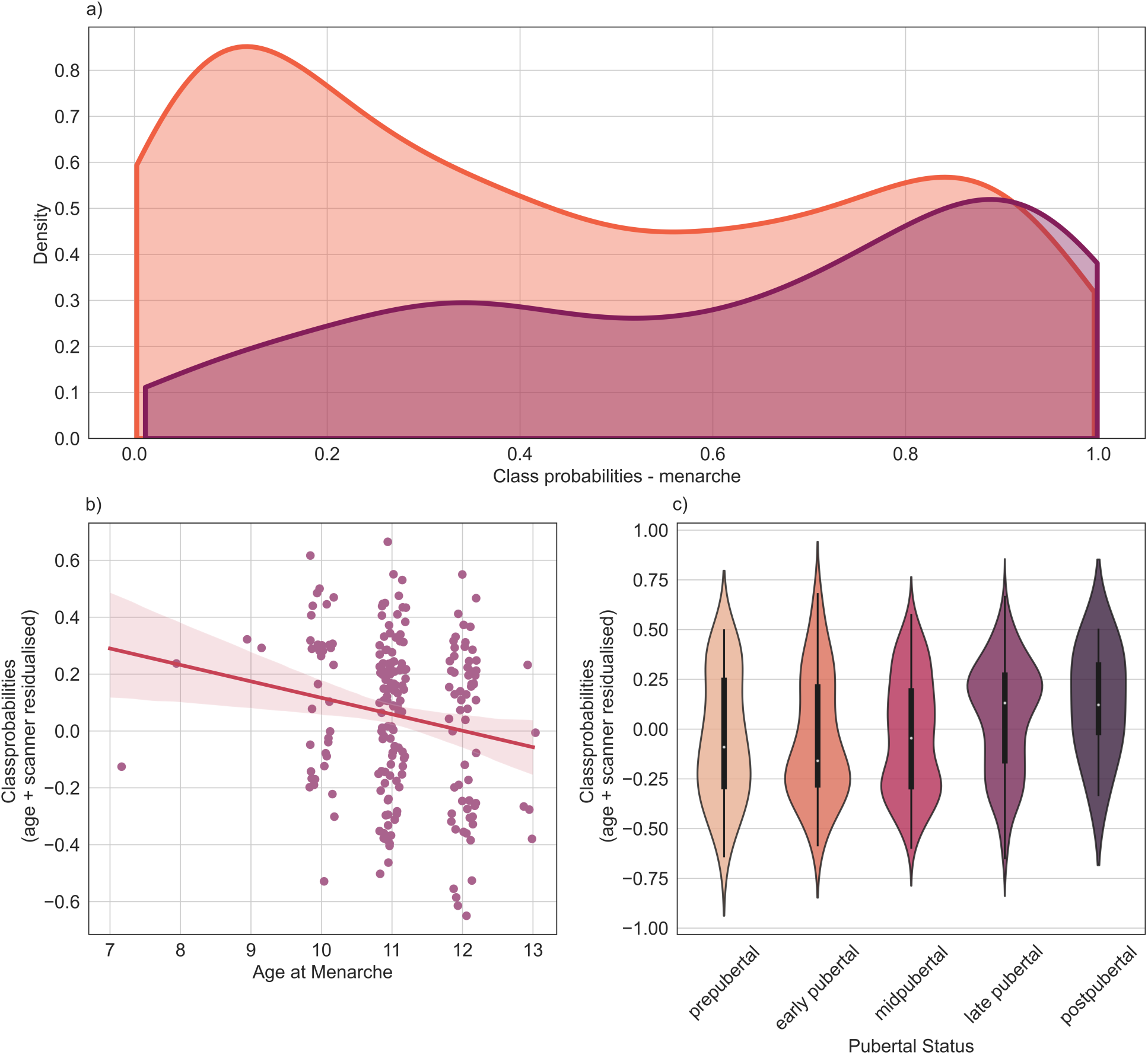
Menarche class probabilities are associated with measures of pubertal timing and status. a) Density plot of post-menarche class probabilities of the pre- and post-menarche groups respectively. Class probability of 1 signifies a 100% confident classification as post-menarche, class probability of 0 signifies a 100% confident classification as pre-menarche. b) Association of age at menarche and menarche class probability controlled for age and scanner. c) Distribution of class probabilities (age- and scanner residualised) by puberty category (youth-reported).

**Figure 3:**
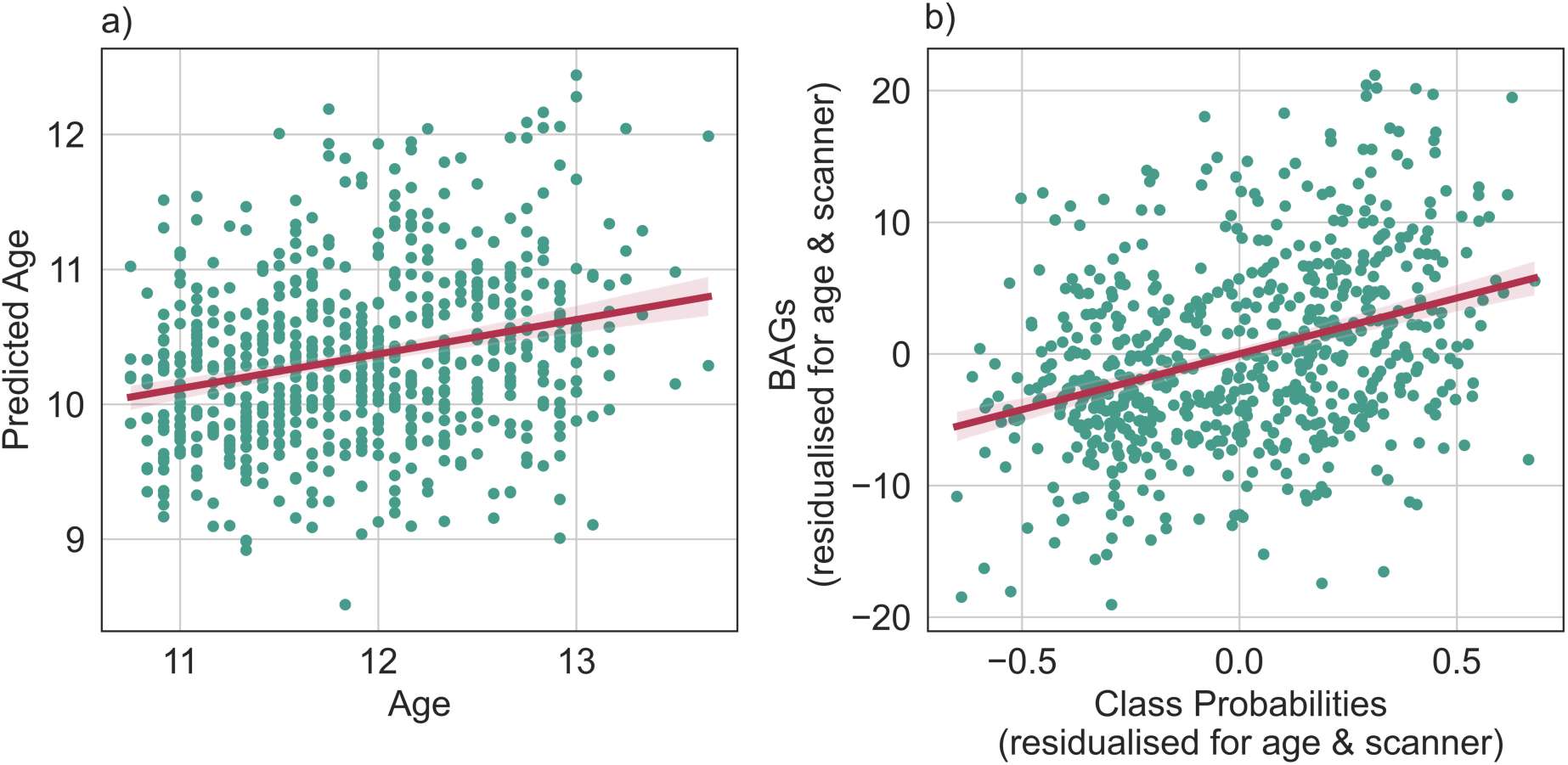
Comparison of menarche classification to a brain age prediction framework. a) Predicted age by age. b) BAGs residualised for age and scanner by menarche class probabilities residualised for age and scanner.

Pubertal development and age are intertwined. Our classifier distinguished between pre- and post-menarcheal females of same age, thereby essentially distinguishing earlier from later pubertal timing relative to age-matched peers. We further sought to investigate whether and to which degree the menarche class probabilities relate to brain age patterns. Our brain age prediction model performed with an RMSE of 2.3 years and a MAE of 1.8 years (5-fold cross validation) in the PNC sample, and permutation tests indicating significant age prediction (p<0.001, 1000 permutations). We next applied the brain age prediction model to the same independent ABCD test sample as used as test sample in the menarche classification. Here, the prediction model performed with an RMSE of 1.8 years and a MAE of 1.6 years (Figure 3a). From the predicted brain ages, we calculated the brain age gap (difference between predicted brain age and chronological age; BAG). These gaps were significantly associated with menarche class probabilities (Fig. 3b), as observed from a linear model controlling for the effect of age and scanner (adj. *R*2 = 0.126, F(1, 648) = 94.51, t = 9.722, p < 0.001). This association stayed significant when including BMI, SES, and race / ethnicity as covariates in the analysis (Supplementary Table S2). BAG was positively associated with pubertal status (youth-reported PDMS: adj. *R*2 = 0.008, F(1, 629) = 5.826, t = 2.414, p = 0.016, caregiver-reported PDMS: adj. *R*2 = 0.009, F(1, 616) = 6.841, t = 2.616, p = 0.009). This effect was descriptively smaller as compared to the class probability effect and also diminshed after including both variables into a single model, in which only class probabilities remained signifcantly associated with pubertal status (t = 3.076, p = 0.002). Interestingly, whereas the menarche class probability was weakly associated with age at menarche as reported above, the brain age gaps were not. Including age at menarche in the model showed no correlation of BAG and age at menarche (adj. *R*2 = 0.366, F(26, 188) = 5.760, t = -1.748, p = 0.082), lending support to the idea that the menarche classification model picks up biological variability additional to that revealed by a brain age model.

## Discussion

The present study aimed at investigating whether structural MRI data can be used to correctly classify pre- vs post-menarche status in adolescent females, thus shedding light on the neurodevelopment associated with pubertal timing. For this, we successfully trained a machine learning model for the classification of pre- and post-menarcheal females in the ABCD cohort while strictly controlling for age-related neurodevelopment through age-matching. To further disentangle age- vs. puberty-related patterns in neurodevelopment, we performed subsequent comparison to a brain age prediction framework that predicts chronological age from MRI, revealing shared and distinct variance in the two machine learning approaches. Finally, we investigated if the class probabilities obtained from menarche classification render a continuous biological marker of pubertal timing that can add relevant information beyond the pre- vs post-menarche dichotomy. Indeed, our results indicate that the probabilities are sensitive to other key variables of pubertal maturation, in particular age at menarche and pubertal status.

### Menarche classification

We argue that leveraging a multivariate, machine learning model helps to integrate information from a collection of brain regions into a single score, which eventually may overcome the inherent complexity of modelling puberty in a univariate fashion and its accompanying statistical considerations [7,44–46] (see [44] for a conceptually similar approach of integrating various sources into a single marker representing pubertal timing). By doing so, our menarche classification model performed with moderate yet significantly above chance accuracy during cross-validation and when applied to a withheld test sample. To rule out that this classification solely mimics a separation of a younger vs. older subgroup of females, we performed a strict age matching prior to model training. Consequently, our results suggest that there is menarche related neuronal variance detectable in structural MRI data. This aligns well with endocrinological trajectories, which are characterized by a substantial, year-long increase in estradiol levels prior to menarche [47] and related findings that estrogens affect neuroplasticity [6,48,49].

### Validation of derived class probabilities

Given the close relationship between pubertal- and age-related neurodevelopment we contrasted the results of our classification model to outcomes of a brain age prediction framework. This approach aimed at exploring the degree to which our derived class probabilities and brain-age estimations capture similar or distinct patterns of neurodevelopmental variation in the adolescent female brain. Testing the brain age prediction model on the above mentioned held-out test sample from the ABCD cohort, we observed highly significant and accurate model performance comparable to results of previous studies modelling brain age in the ABCD cohort [6]. Our derived menarche class probabilities were positively related to brain age gaps (BAGs, i.e., the difference between someone’s brain and chronological age), matching earlier results that associated brain age with pubertal status (e.g., [6]) and pubertal timing (e.g., [44]). Individuals with higher class probabilities (i.e., a higher probability of being classified as post-menarche) also had higher brain-age gaps (i.e., an indication of a more mature brain in relation to their chronological ages), suggesting that both approaches capture variations in adolescent brain development related to advanced brain maturation. Our work however extends previous findings, by showing that our menarche classification approach seems to be able to better exploit traces of pubertal timing in the brain that are specific to puberty and go beyond the traces of age-related neurodevelopment that are captured by a brain age prediction framework. This finding is in line with previous suggestions that puberty related processes may be a more sensitive measure to investigate adolescent brain development compared to age-related neurodevelopment [6–9].

To prove the additional benefit of studying neurodevelopment from a puberty-focused perspective and to further substantiate the biological validity and capability of the class probabilities of capturing meaningful biological variance, we furthermore aimed at investigating the probabilities associations with puberty-related measurements. In the ABCD study, puberty is assessed by different means, ranging from hormonal measurements to self-reported evaluation of perceived pubertal maturation (see [50]. The latter allows to localize individuals in different pubertal stages or categories ranging from pre- to post-pubertal. As described before, for females the score is derived by summing over ratings of key physical changes, such as breast development and pubic hair growth, but also the (non-) completion of menarche [14]. Since menarche is directly incorporated in the pubertal category scores, we additionally showed that higher pubertal category scores (thus indicating a later pubertal stage) are associated with higher class probabilities, which serves as an important sanity check for our proposed approach. Higher BAGs were also significantly associated with higher pubertal category scores, however with a descriptively smaller effect sized compared to the class probabilities. Furthermore, after including both variables into a model, only the effect of class probabilities remained significant. Furthermore, we show that post-menarche class probabilities were weakly yet significantly associated with age at menarche. In contrast, the association between BAG and age at menarche was not significant. This suggests that there are traces of pubertal timing in the brain that go beyond patterns of age-related brain development, and that these traces can be more successfully exploited by our proposed menarche classification model than by the brain age prediction framework. Interestingly, the pattern of higher class probabilities in females that underwent early or earlier menarche, resonates with the hypothesis that the brain might be more susceptible to the hormonal influences of puberty at a younger age and that, therefore, individuals who experience an earlier menarche undergo the increase of gonadal hormones at a time when their brain is relatively more sensitive to their effects on neuroplasticity [51,52]. Of note, after adding SES, race/ethnicity, and BMI as covariates into our analyses, the association between the class probabilities and age at menarche diminished. This observation matches previous reports about the close link between pubertal processes, for example pubertal timing (e.g., operationalized as age at menarche) [53] and these covariates (e.g., [54–56]). Importantly, associations between the aforementioned covariates and puberty were also replicated for ABCD 2-year follow up data, which we used in the current study [14]. While we consider it important to understand the associations with these covariates, their interplay is difficult to disentangle with the data at hand, given the relationship between pubertal timing and these variables. We argue that these results rather warrant further systematic investigation of the interplay between all factors in the equation.

### Methodological considerations and future directions

Potential limitations may stem from the fact that we limited our machine learning model to structural imaging data from cortical and subcortical regions. While this decision resonates with well-replicated findings of cortical and subcortical grey matter changes during puberty [7], integrating additional imaging features, such as white matter measures, may help in a more holistic investigation of pubertal timing. While myelination plays a crucial role in shaping the human brain during adolescence, findings regarding pubertal maturation appear to be either mixed regarding different measures of white matter (e.g., [5,7]) or lack previous investigations. Furthermore, while we followed a common approach of training our brain-age prediction model in an independent dataset (see e.g., [6,42]) and applying it to our target sample in the ABCD cohort, recent work from Ray and colleagues [57] suggests that refined brain age models (i.e., a combination of pre-trained models with subsequent finetuning on a fraction of the target data) may improve model performance and thus also downstream analyses. Lastly, our work focuses on a proof of concept on the 2-year follow up data of the ABCD study. With additional longitudinal data becoming available through upcoming releases, the ABCD study depicts an unprecedented resource to validate our model and proof its usability, since more and more female will eventually undergo their menarche.

## Perspective and Significance

This work may be seen as a proof of principle that pubertal timing can be classified from brain imaging data. Previous studies that have used age focused approaches like brain age prediction frameworks have found associations with pubertal measures [44], yet our results suggest that grounding the modelling in puberty data directly may yield brain based markers that are even more sensitive to pubertal status and timing. Future studies may thus further explore similar approaches toward the development of brain-based puberty markers that may be useful in downstream analyses in developmental neuroscience.

## Conclusion

We introduced a machine learning approach that classifies menarche status of adolescent females from their cortical and subcortical structural MRI data. The derived continuous brain-based class probabilities captured shared but also unique variations of adolescent neurodevelopment when compared to a brain-age prediction model. Taken together, our results suggest that there are markers of menarche in the brain that can be formalized into a continuous class probability, which might in the future be developed toward an objective brain-based marker of pubertal timing.

## Data Availability

Data used in this study has been shared with the authors under the data usage agreements of the respective cohorts (i.e., ABCD and PNC). To use these resources, interested scholars can obtain access by signing their own respective data usage agreements with the officials. Code used to analyze the data will be made available on github upon acceptance of the manuscript.

## Declarations

### Ethics approval

PNC study procedures were approved by institutional review boards of the University of Pennsylvania and the Children’s Hospital of Philadelphia. All participants or their caregiver provided written informed consent. ABCD procedures were approved by either the local site Institutional Review Board or by local Institutional Review Board reliance agreements with the central Institutional Review Board at the University of California. All participants and their parents provided written informed consent.

### Consent for publication

All subjects consented to study inclusion as part of the data acquisition in the ABCD or PNC cohort. Data was shared with the authors via dedicated data use agreements.

### Author contribution

NG had the initial idea to build a menarche classifier. NG and TK conceptualized the study. TK processed the imaging data. DK curated the ABCD data. NG analysed the data. NG visualized the data. All authors interpreted the data and wrote the paper. TK received funding.

## Acknowledgement

The authors used data from the Philadelphia Neurodevelopmental Cohort (PNC, https://www.ncbi.nlm.nih.gov/projects/gap/cgi-bin/study.cgi?study_id=phs000607.v3.p2, access permission no 29782) and the Adolescent Brain Cognitive Development^SM^ Study (ABCD, abcdstudy.org), Support for the collection of the PNC data set was provided by grant #RC2MH089983 awarded to Raquel Gur, MD, PhD, and #RC2MH089924 awarded to Hakon Hakonarson, MD, PhD. ABCD data, held in the NIMH Data Archive (NDA), is a multisite, longitudinal study designed to recruit more than 10,000 children age 9-10 and follow them over 10 years into early adulthood. The ABCD Study® is supported by the National Institutes of Health and additional federal partners under award numbers U01DA041048, U01DA050989, U01DA051016, U01DA041022, U01DA051018, U01DA051037, U01DA050987, U01DA041174, U01DA041106, U01DA041117, U01DA041028, U01DA041134, U01DA050988, U01DA051039, U01DA041156, U01DA041025, U01DA041120, U01DA051038, U01DA041148, U01DA041093, U01DA041089, U24DA041123, U24DA041147. A full list of supporters is available at https://abcdstudy.org/federal-partners.html. A listing of participating sites and a complete listing of the study investigators can be found at https://abcdstudy.org/consortium_members/. PNC and ABCD consortium investigators designed and implemented the respective studies and/or provided data but did not participate in the analysis or writing of this report. This manuscript reflects the views of the authors and does not necessarily reflect the opinions or views of any other agency, organization, employer or company.

## Supplementary Information

<u>Additional File 1</u>: Supplementary Table S1, S2

