## Supplementary File for "Menarche, pubertal timing and the brain: female-specific patterns of brain maturation beyond age-related development"

Postal address:
Department of Psychiatry and Psychotherapy, Calwerstr. 14, 72076 Tübingen

### Additional File 1

Supplementary Tables

*Supplementary Table S1: Cortical and subcortical features for menarche classification and brain age prediction.*

| Thickness | Volume | Area | SubcorticalVolumes |
|---|---|---|---|
| **bankssts_thicknessstd** | bankssts_volume | bankssts_area | smri_vol_scs_ltventriclelh |
| **caudalanteriorcingulate_thicknessstd** | caudalanteriorcingulate_volume | caudalanteriorcingulate_area | smri_vol_scs_inflatventlh |
| **caudalmiddlefrontal_thicknessstd** | caudalmiddlefrontal_volume | caudalmiddlefrontal_area | smri_vol_scs_crbwmatterlh |
| **cuneus_thicknessstd** | cuneus_volume | cuneus_area | smri_vol_scs_crbcortexlh |
| **entorhinal_thicknessstd** | entorhinal_volume | entorhinal_area | smri_vol_scs_tplh |
| **fusiform_thicknessstd** | fusiform_volume | fusiform_area | smri_vol_scs_caudatelh |
| **inferiorparietal_thicknessstd** | inferiorparietal_volume | inferiorparietal_area | smri_vol_scs_putamenlh |
| **inferiortemporal_thicknessstd** | inferiortemporal_volume | inferiortemporal_area | smri_vol_scs_pallidumlh |
| **isthmuscingulate_thicknessstd** | isthmuscingulate_volume | isthmuscingulate_area | smri_vol_scs_3rdventricle |
| **lateraloccipital_thicknessstd** | lateraloccipital_volume | lateraloccipital_area | smri_vol_scs_4thventricle |
| **lateralorbitofrontal_thicknessstd** | lateralorbitofrontal_volume | lateralorbitofrontal_area | smri_vol_scs_bstem |
| **lingual_thicknessstd** | lingual_volume | lingual_area | smri_vol_scs_hpuslh |
| **medialorbitofrontal_thicknessstd** | medialorbitofrontal_volume | medialorbitofrontal_area | smri_vol_scs_amygdalalh |
| **middletemporal_thicknessstd** | middletemporal_volume | middletemporal_area | smri_vol_scs_aal |
| **parahippocampal_thicknessstd** | parahippocampal_volume | parahippocampal_area | smri_vol_scs_ltventriclerh |
| **paracentral_thicknessstd** | paracentral_volume | paracentral_area | smri_vol_scs_inflatventrh |
| **parsopercularis_thicknessstd** | parsopercularis_volume | parsopercularis_area | smri_vol_scs_crbwmatterrh |
| **parsorbitalis_thicknessstd** | parsorbitalis_volume | parsorbitalis_area | smri_vol_scs_crbcortexrh |
| **parstriangularis_thicknessstd** | parstriangularis_volume | parstriangularis_area | smri_vol_scs_tprh |
| **pericalcarine_thicknessstd** | pericalcarine_volume | pericalcarine_area | smri_vol_scs_caudaterh |
| **postcentral_thicknessstd** | postcentral_volume | postcentral_area | smri_vol_scs_putamenrh |
| **posteriorcingulate_thicknessstd** | posteriorcingulate_volume | posteriorcingulate_area | smri_vol_scs_pallidumrh |
| **precentral_thicknessstd** | precentral_volume | precentral_area | smri_vol_scs_hpusrh |
| **precuneus_thicknessstd** | precuneus_volume | precuneus_area | smri_vol_scs_amygdalarh |
| **rostralanteriorcingulate_thicknessstd** | rostralanteriorcingulate_volume | rostralanteriorcingulate_area | smri_vol_scs_aar |
| **rostralmiddlefrontal_thicknessstd** | rostralmiddlefrontal_volume | rostralmiddlefrontal_area | smri_vol_scs_ccps |
| **superiorfrontal_thicknessstd** | superiorfrontal_volume | superiorfrontal_area | smri_vol_scs_ccmidps |
| **superiorparietal_thicknessstd** | superiorparietal_volume | superiorparietal_area | smri_vol_scs_ccct |
| **superiortemporal_thicknessstd** | superiortemporal_volume | superiortemporal_area | smri_vol_scs_ccmidat |
| **supramarginal_thicknessstd** | supramarginal_volume | supramarginal_area | smri_vol_scs_ccat |
| **frontalpole_thicknessstd** | frontalpole_volume | frontalpole_area | |
| **temporalpole_thicknessstd** | temporalpole_volume | temporalpole_area | |
| **transversetemporal_thicknessstd** | transversetemporal_volume | transversetemporal_area | |
| **insula_thicknessstd** | insula_volume | insula_area | |

Note: Naming refers to labels obtained in Freesurfer 7 in the PNC cohort. For the ABCD study, we used the same features, yet some may have been labelled differently in the ABCD data base.

*Supplementary Table S2: Results of association analyses when including BMI, SES and race / ethnicity as covariates.*

| | **variable** | **t-value** | **p-value** |
|---|---|---|---|
| Menarche class probabilities ~ age at menarche + age + scanner$_{dummy\text{-}variables}$ + BAG$_{residualised}$ + BMI + SES + race / ethnicity + constant | | | |
| | age at menarche | -0.946 | 0.634 |
| | age | 3.774 | < 0.001 |
| | BAG $_{residualised}$ | 3.637 | < 0.001 |
| | BMI | 0.657 | 0.512 |
| | SES | -0.447 | 0.156 |
| | race/ethnicity $_{asian}$ | -1.218 | 0.225 |
| | race/ethnicity $_{black}$ | -0.902 | 0.369 |
| | race/ethnicity $_{hispanic}$ | -1.485 | 0.140 |
| | race/ethnicity $_{white}$ | -2.998 | 0.003 |
| | race/ethnicity $_{other}$ | -2.697 | 0.008 |
| | constant | -2.340 | 0.021 |
| Pubertal Status$_{(c/y)}$ ~ menarche class probabilities$_{residualised}$ + BAG$_{residualised}$ + BMI + SES + race / ethnicity + constant | | | |
| Caregiver-reported | menarche class probabilities $_{residualised}$ | 0.484 | 0.629 |
| | BAG $_{residualised}$ | 1.321 | 0.187 |
| | BMI | 8.479 | < 0.001 |
| | SES | -1.053 | 0.293 |
| | race/ethnicity $_{asian}$ | 2.040 | 0.042 |
| | race/ethnicity $_{black}$ | 3.917 | < 0.001 |
| | race/ethnicity $_{hispanic}$ | 3.896 | < 0.001 |
| | race/ethnicity $_{white}$ | 2.071 | 0.039 |
| | race/ethnicity $_{other}$ | 2.592 | 0.010 |
| | constant | 11.374 | < 0.001 |
| Youth-reported | menarche class probabilities $_{residualised}$ | 0.625 | 0.532 |
| | BAG $_{residualised}$ | 1.267 | 0.206 |
| | BMI | 6.833 | < 0.001 |
| | SES | -0.151 | 0.880 |
| | race/ethnicity $_{asian}$ | 1.664 | 0.097 |
| | race/ethnicity $_{black}$ | 3.032 | 0.003 |
| | race/ethnicity $_{hispanic}$ | 4.767 | < 0.001 |
| | race/ethnicity $_{white}$ | 2.644 | 0.008 |
| | race/ethnicity $_{other}$ | 3.605 | < 0.001 |
| | constant | 11.608 | < 0.001 |

Note: BAG = Brain Age Gap (predicted age – chronological age), BMI = Body Mass Index, SES = Socioeconomic Status, BAG$_{residualised}$ = residualised BAG score produced by regressing age and scanning site on BAG, menarche class probabilities$_{residualised}$ = residualised menarche class probabilities produced by regressing age and scanning site on menarche class probabilities, Pubertal Status$_{(c/y)}$ = pubertal status category reported bei either caregiver or youth. MRI scanner dummy variables were omitted from the table for legibility but were included in the modelling. 1 out of 26 scanning devices was significantly associated with menarche class probability (HASH4036a433, t = -2.593, p = 0.011), reflecting scan site specific population characteristics.
